# Deformation prior-guided deep learning for post hoc motion deblurring in single-phase cardiac computed tomography

**DOI:** 10.64898/2026.11.25.26368675

**Authors:** Tatsuya Nishii, Rina Sakai, Tatsuya Saito, Toshimitsu Tanaka, Kensuke Umehara, Junko Ota, Takuma Kobayashi, Atsushi K. Kono, Yasutoshi Ohta, Tetsuya Fukuda

## Abstract

**Purpose:** To develop and evaluate DeTangle, a deep learning-based method for cardiac computed tomography (CT) deblurring using internally estimated deformation-magnitude maps and single-phase reconstructed images.

**Materials and Methods:** This retrospective study included 60 patients (median age, 61 years; 40 males) who underwent cardiac CT between January 2024 and October 2025. Patients comprised training, validation, and internal test sets (30, five, and 25 patients, respectively). DeTangle jointly optimizes deformation-map estimation and restoration using privileged deformation information during training but requires only target-phase reconstructed images at inference. Full reconstructions (250 ms) served as inputs and half reconstructions (66 ms) as references. The structural similarity index measure (SSIM) was assessed in 424 test phases. DeTangle and the image-only Baseline were compared using five-point visual scores in 200 phases and edge measurements in 100 phases. Patient-clustered generalized estimating equations incorporated Holm adjustment.

**Results:** Mean SSIM (± SD) was higher with DeTangle than Baseline (0.972 ± 0.006 vs 0.968 ± 0.008, P < .001). Mean coronary and left ventricular myocardial scores were also higher (3.3 ± 1.0 vs 2.9 ± 1.1 and 4.2 ± 1.0 vs 3.5 ± 1.3, respectively; both P < .001). DeTangle increased edge rise slope and reduced edge rise distance at the interventricular septum and right and left coronary arteries (all P < .05). At least one primary visual score was lower with DeTangle than with Baseline in nine of 200 phases (4.5%).

**Conclusion:** DeTangle improved reference-image agreement, coronary and myocardial visual scores, and edge sharpness versus Baseline, although some phases showed localized image-quality deterioration.

## Introduction

Cardiac motion can impair delineation of the coronary arteries, myocardium, and valvular structures. Faster gantry rotation and dual-source computed tomography (CT) have improved temporal resolution [1] and coronary evaluability [2]. High temporal resolution remains important for coronary image quality, even in photon-counting CT [3]. However, mechanical constraints limit further increases in rotation speed, necessitating motion compensation for residual blur [4]. Motion-correction methods incorporate estimated motion fields into projection-based reconstruction [4–7], whereas others deform images using motion estimated from adjacent cardiac phases [8]. Such methods improve visualization of coronary arteries [5,6,9,10], the aortic annulus[8], and mitral valves [11].

Deep learning-based postprocessing operates on reconstructed images without projection data. Applications include denoising coronary CT angiography [12], myocardial delayed-enhancement images[13], and improving spatial resolution [14]. Motion artifact reduction using direct image translation or multiple cardiac phases has also been investigated [15–17]. However, only single-phase reconstructed images may be available for postprocessing.

Motion-guided reconstruction [4–7] and multiphase deep learning [16,17] support using motion information for deblurring. We hypothesized that deformation maps derived from paired reconstructions could serve as privileged information during training to improve restoration from single-phase images. We designed DeTangle with the aim of learning to disentangle motion effects from anatomy. To implement this concept, we jointly trained one network to estimate a deformation-magnitude map from three adjacent slices of a single cardiac phase and another to use that map to guide restoration of the central slice. The purpose of this study was to develop and evaluate DeTangle for reducing motion blur in coronary and noncoronary structures on cardiac CT.

## Materials and Methods

### 1. Study design and patients

The Research Ethics Committee of the National Cerebral and Cardiovascular Center approved this retrospective, single-center study (R20112-2) and waived written informed consent. Consecutive patients with structural heart disease who underwent full-cycle cardiac CT on a dual-source scanner (SOMATOM Force; Siemens Healthineers) between January 2024 and October 2025 were eligible. Exclusion criteria included arrhythmia with a heart rate variation ≥5 beats/min during scanning, severe artifacts from inadequate breath-holding, and unavailable projection data (Figure 1). Sample size was feasibility-based, and data were de-identified. Patients were assigned chronologically to non-overlapping training, validation, and internal test sets of 30, five, and 25 patients, respectively.

**Figure 1.**
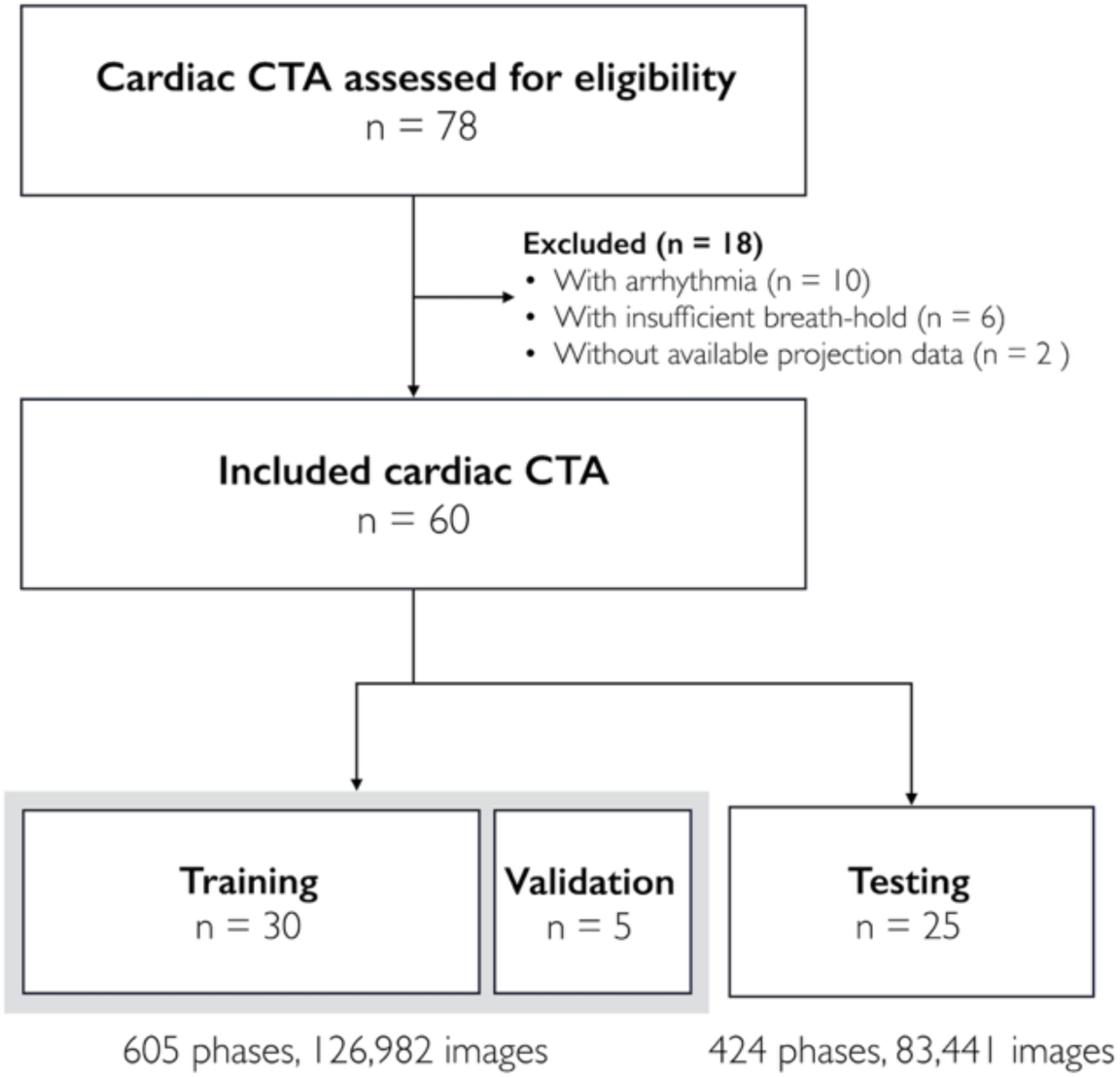
Study flowchart. CTA, computed tomography angiography.

### 2. Image acquisition and preparation

Retrospective electrocardiogram (ECG)-gated helical CT was performed without ECG-based tube current modulation in “Chest pain” scan mode, yielding motion-blurred full reconstructions (MB-Full) and motion-reduced half reconstructions (MR-Half). Reconstruction start times were spaced at 50-ms intervals. Each MB-Full image, reconstructed over a 250-ms window, served as the model input. Within each window, early (MR-Half₁, 0–66 ms) and late (MR-Half₂, 150–216 ms) half reconstructions were generated at the highest available temporal resolution. These times are relative to the start of the corresponding MB-Full window, not to the ECG R wave. Both half reconstructions underwent post hoc deep learning denoising [12,18,19] to reduce noise differences associated with their shorter reconstruction windows; MB-Full was not denoised. Denoised MR-Half₁ served as the restoration reference for the corresponding MB-Full image. To derive displacement-magnitude maps (RealDM), we registered denoised MR-Half₂ to denoised MR-Half₁ using symmetric normalization (SyN)[20] in ANTsPy [21]. RealDM was used for model development and test-phase motion stratification. Acquisition parameters and processing details are provided in Supplemental Table S1 and Methods S1.

### 3. Model development and inference

DeTangle (<u>De</u>blurring te<u>a</u>cher-distilled <u>n</u>etwork levera<u>g</u>ing privi<u>le</u>ged deformation priors) uses two Swin-Conv-UNet-based networks (SCUNet) [22] (Figure 2 and Supplemental Figure S1). DMNet estimates a deformation-magnitude map (cDM) from the target MB-Full slice and two adjacent slices. Deblur uses these slices and cDM to restore the target slice. We compared four training configurations (Supplemental Table S2). Baseline used images alone; learning using privileged information (LUPI)-only used image-only restoration with distillation from Teacher; DeTangle jointly optimized DMNet and Deblur with deformation-map supervision and output distillation; and Teacher used registration-derived RealDM directly. At inference, DeTangle requires only MB-Full images. Training details are provided in Supplemental Methods S2.

**Figure 2.**
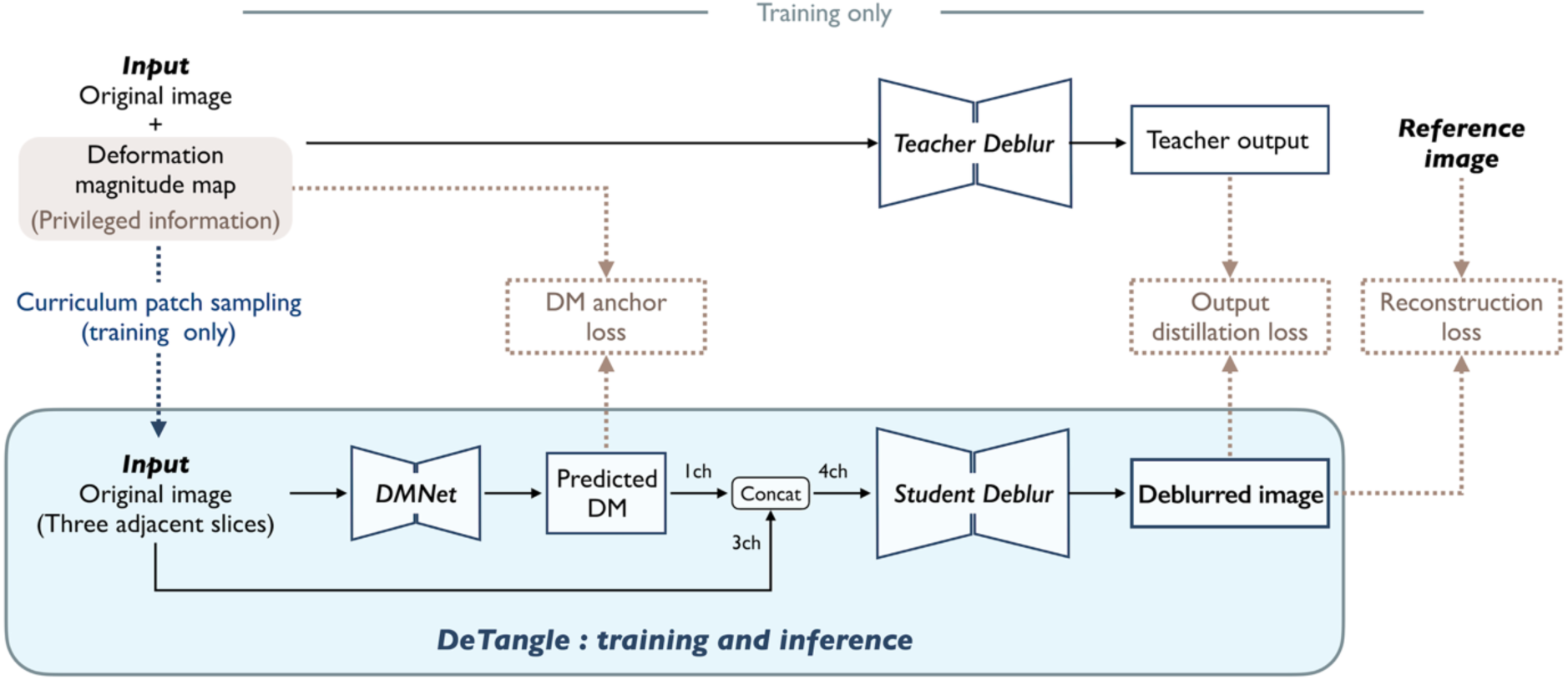
DeTangle training and inference framework. DeTangle comprises two Swin-Conv-UNet (SCUNet)-based networks: DMNet, which estimates deformation magnitude, and Student Deblur, which restores the target image. DMNet estimates a deformation-magnitude map (cDM) from three adjacent motion-blurred full-reconstruction CT slices (MB-Full). After alignment, cDM and the three slices form a four-channel input to Student Deblur. During training, registration-derived deformation-magnitude maps (RealDM) provide privileged input to Teacher Deblur, supervise cDM through DM anchor loss, and guide curriculum-based training-patch sampling. Student Deblur is supervised by motion-reduced half-reconstruction reference images (MR-Half) and Teacher output through reconstruction loss and output distillation, respectively. The light blue region indicates the DeTangle pathway shared by training and inference. At inference, only DMNet and Student Deblur are used, requiring MB-Full images alone. Black solid arrows: forward computation; brown dotted arrows: training-loss calculations; blue dotted arrows: curriculum patch-sampling guidance. DM, deformation magnitude map; Concat, channel-wise concatenation; ch, channel; CT, computed tomography.

### 4. Image quality evaluation

Reference agreement was evaluated across all 424 phases from the 25-patient internal test set. Reader 1 evaluated 200 phases selected across systolic, mid-diastolic, and out-of-window periods. Reader 2 and quantitative analyses used a 100-phase subset (Supplemental Methods S3).

#### 1.1. Agreement with reference images

Agreement with MR-Half was assessed using the structural similarity index measure (SSIM), peak signal-to-noise ratio (PSNR), and learned perceptual image patch similarity (LPIPS). We quantified phase-level motion using the 95th percentile of RealDM values across slices. Motion was stratified into tertiles (Supplemental Methods S1.3). Tertiles of these scores across all 424 test phases defined low (<2.6 mm), intermediate (2.6 to <4.2 mm), and high (≥4.2 mm) motion.

#### 1.2. Visual image quality

Readers independently scored Baseline and DeTangle images presented in randomized, model-blinded order. Reader 1 (a board-certified radiologist) scored all 200 phases, and Reader 2 (a board-certified radiological technologist) scored the 100-phase subset, each assessing nine regions on a five-point scale. The principal outcomes were three-vessel mean coronary and left ventricular myocardial scores. Scoring criteria are detailed in Supplemental Methods S4.

#### 1.3. Quantitative image quality

Reader 1 placed image-specific regions of interest (ROIs) on MB-Full, MR-Half, Baseline, and DeTangle images in the 100-phase subset. The edge rise slope (ERS) and edge rise distance (ERD) were measured at the interventricular septum, right coronary artery (RCA), and left coronary artery (LCA). ERS and ERD quantified edge sharpness over the 10%–90% intensity transition. CT attenuation, image noise, signal-to-noise ratio (SNR), and contrast-to-noise ratio (CNR) were also evaluated. ROI placement and measurement procedures are described in Supplemental Methods S5.

#### 4.4. Exploratory application to half reconstructions

DeTangle was also applied to denoised MR-Half in the 100-phase subset, with ERD and ERS compared against MR-Half and DeTangle-processed MB-Full (Supplemental Methods S6).

### 5. DM input experiments and inference time

We evaluated the effects of RealDM perturbation and partial or complete cDM substitution on SSIM, with network weights fixed (Supplemental Methods S7). Inference time per slice was calculated from total test-set processing time on a GeForce RTX 4070 Ti SUPER using automatic mixed precision.

### 6. Statistical analysis

Patient characteristics were summarized as median (interquartile range [IQR]) or count (%), and phase metrics as mean ± standard deviation (SD). Generalized estimating equations (GEEs) used patient clusters, an independent working correlation, and a Mancl–DeRouen bias-reduced sandwich variance estimator. Analyses were performed using R (version 4.6.0, R Foundation for Statistical Computing); P < .05 indicated statistical significance.

Reference agreement and quantitative outcomes were analyzed using Gaussian GEEs with Holm-adjusted pairwise comparisons. Motion-dependent trends were assessed using linear trends or image-type × ordered-motion-group interactions. Visual outcomes were analyzed using Gaussian GEEs for coronary mean scores and cumulative logit ordinal GEEs for myocardial and regional scores, with Holm adjustment. Interreader agreement was assessed using linearly weighted Cohen κ. Details of multiple-comparison adjustments, exploratory perturbation and mixing analyses, and confidence interval estimation are provided in Supplemental Methods S8.

## Results

Of 78 patients screened, 60 were included (Figure 1). The median age was 61 years (IQR, 55–72 years), 40 patients (67%) were male, and the median heart rate was 60 beats/min (IQR, 56–67 beats/min) (Table 1). The training, validation, and test sets comprised 521, 84, and 424 cardiac phase volumes, respectively. Low/intermediate/high motion-group counts were 130/149/145 for all test phases, 71/74/55 for visual assessment, and 42/37/21 for quantitative assessment. Eight phases from one patient were excluded from distal left anterior descending coronary artery scoring, leaving 192 phases. All other measurements and scores were available.

**Table 1.** Patient characteristics and cardiac computed tomography acquisition parameters.

| **Variables** | **Total** | **Training and validation** | **Test** |
|---|---|---|---|
|  | **(N = 60)** | **(N = 35)** | **(N = 25)** |
| Age, years | 61 (55, 72) | 62 (55, 73) | 60 (51, 70) |
| Sex, Female | 20 (33) | 10 (29) | 10 (40) |
| Male | 40 (67) | 25 (71) | 15 (60) |
| Body mass index, kg/m<sup>2</sup> | 22.7 (20.2, 25.8) | 21.1 (19.7, 25.8) | 23.5 (21.6, 25.9) |
| Heart rate, bpm | 60 (56, 67) | 60 (54, 65) | 61 (57, 68) |
| RR interval, msec | 1000 (902, 1071) | 1000 (930, 1112) | 984 (882, 1053) |
| Premedication for scan |  |  |  |
| Nitroglycerin | 33 (55) | 21 (60) | 12 (48) |
| Beta-blocker | 18 (30) | 10 (29) | 8 (32) |
| Contrast materials |  |  |  |
| 370 mgI/mL | 27 (45) | 16 (46) | 11 (44) |
| 320 mgI/mL | 33 (55) | 19 (54) | 14 (56) |
| Injection rate, mL/sec | 4.6 (4.3, 4.8) | 4.6 (4.2, 4.8) | 4.7 (4.3, 5.1) |
| Total contrast amounts, mL | 74.7 (68.4, 77.9) | 72.0 (64.4, 76.8) | 76.5 (70.1, 80.4) |
| Tube potential |  |  |  |
| 70 kV | 14 (23) | 10 (29) | 4 (16) |
| 80 kV | 28 (47) | 15 (43) | 13 (52) |
| 90 kV | 13 (22) | 8 (23) | 5 (20) |
| 100 kV | 3 (5) | 2 (6) | 1 (4) |
| 120 kV | 2 (3) | 0 (0) | 2 (8) |
| CT dose index, mGy | 47.0 (35.6, 63.7) | 46.5 (34.2, 63.1) | 48.8 (36.2, 64.3) |
| Dose–length product, mGy·cm | 840.9 (625.9, 1092.4) | 831.7 (604.5, 1092.5) | 850.0 (643.1, 1052.0) |

Continuous variables are reported as median (25th, 75th percentiles), and categorical variables as number of patients (percentage within each cohort).
CT, computed tomography

### Reference agreement

Mean SSIM relative to MR-Half was 0.968 ± 0.008 for Baseline, 0.969 ± 0.007 for LUPI-only, 0.972 ± 0.006 for DeTangle, and 0.976 ± 0.006 for Teacher (Figure 3). DeTangle outperformed Baseline and LUPI-only in SSIM, PSNR, and LPIPS but remained inferior to Teacher (all Holm-adjusted P < .001; Table 2). Unprocessed MB-Full had a mean SSIM of 0.960 ± 0.010. Validation SSIM curves over 210 training epochs for Baseline and DeTangle are shown in Supplemental Figure S2. Representative patients are shown in Figures 4 and 5.

**Figure 3.**
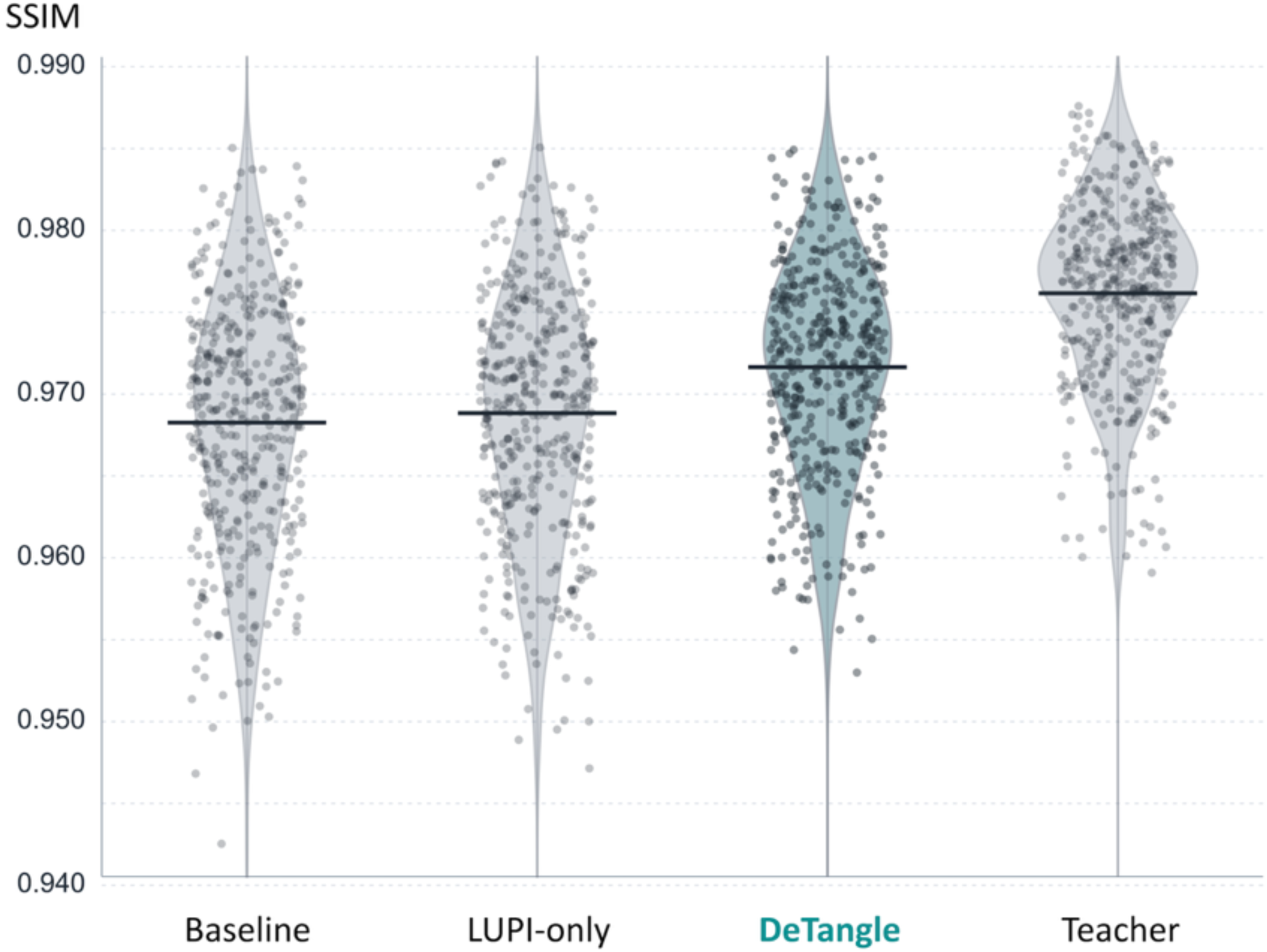
Structural similarity to reference images across four models. Bean plots show phase-level structural similarity index (SSIM) relative to motion-reduced half-reconstruction reference images (MR-Half). Each dot represents a cardiac phase, and horizontal bars indicate means. Mean ± SD are 0.968 ± 0.008 (Baseline), 0.969 ± 0.007 (LUPI-only), 0.972 ± 0.006 (DeTangle), and 0.976 ± 0.006 (Teacher). All six pairwise SSIM comparisons are statistically significant (all Holm-adjusted P < .001). LUPI, learning using privileged information.

**Table 2.** PSNR and LPIPS relative to MR-Half by model.

| **Model** | **PSNR (dB)** | **LPIPS** |
|---|---|---|
| Baseline | 37.79 ± 1.61 | 0.05928 ± 0.01938 |
| LUPI-only | 37.85 ± 1.57 | 0.05926 ± 0.01910 |
| DeTangle | 38.53 ± 1.37 | 0.0560 ± 0.0183 |
| Teacher | 39.57 ± 1.31 | 0.0523 ± 0.0173 |

Values are mean ± standard deviation. The motion-reduced half reconstruction served as the reference.
\* For both PSNR and LPIPS, DeTangle differed significantly from Baseline, LUPI-only, and Teacher (all two-sided Holm-adjusted *P* <.001). P values were adjusted across all six pairwise model comparisons within each metric.
PSNR, peak signal-to-noise ratio; LPIPS, learned perceptual image patch similarity.

**Figure 4.**
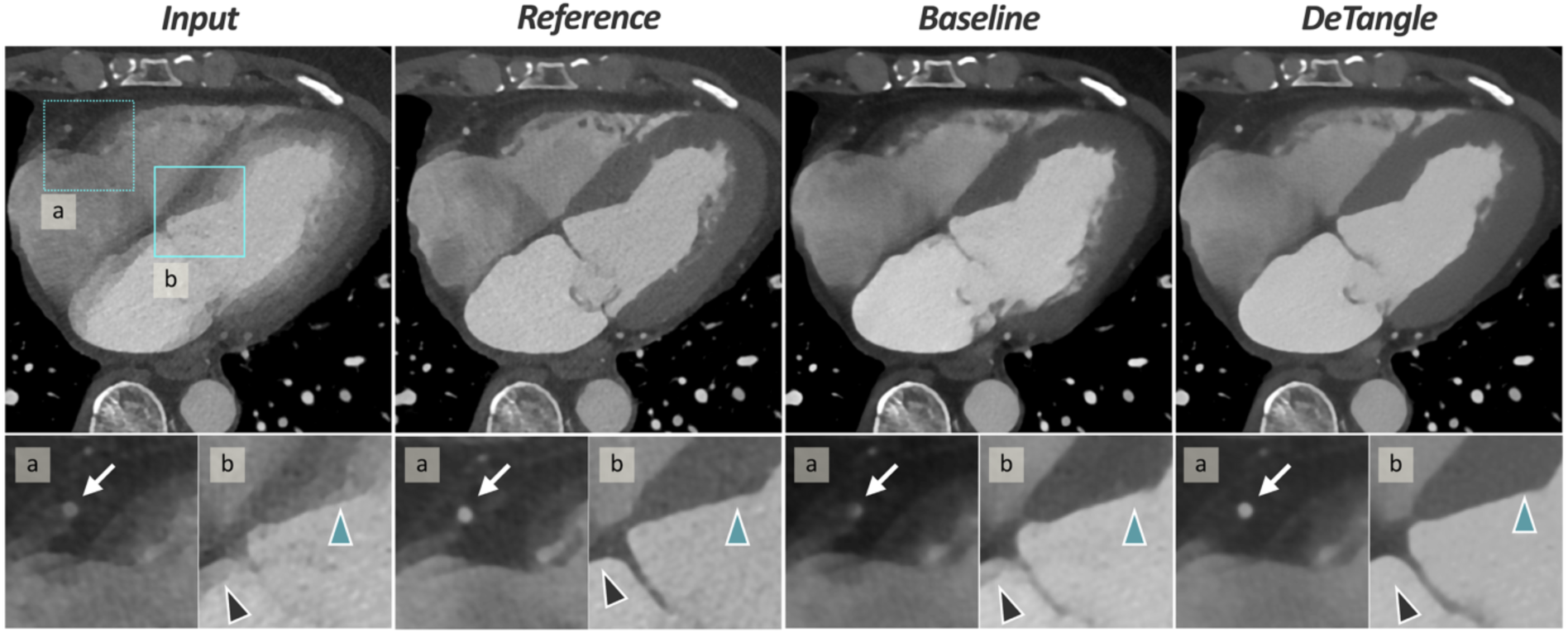
Representative case of improved cardiac computed tomography image quality with DeTangle. End-systolic cardiac computed tomography (CT) images with reconstruction starting 300 ms after the R wave of a man in his 50s show motion-blurred full reconstruction (MB-Full), motion-reduced half reconstruction (MR-Half), Baseline, and DeTangle (left to right), with magnified regions below. (a) The right coronary artery (white arrows) remains blurred after Baseline processing and is better delineated with DeTangle, resembling the reference. (b) Baseline improves delineation of the interventricular septum (blue arrowheads). DeTangle improves delineation of the interatrial septum (black arrowheads).

**Figure 5.**
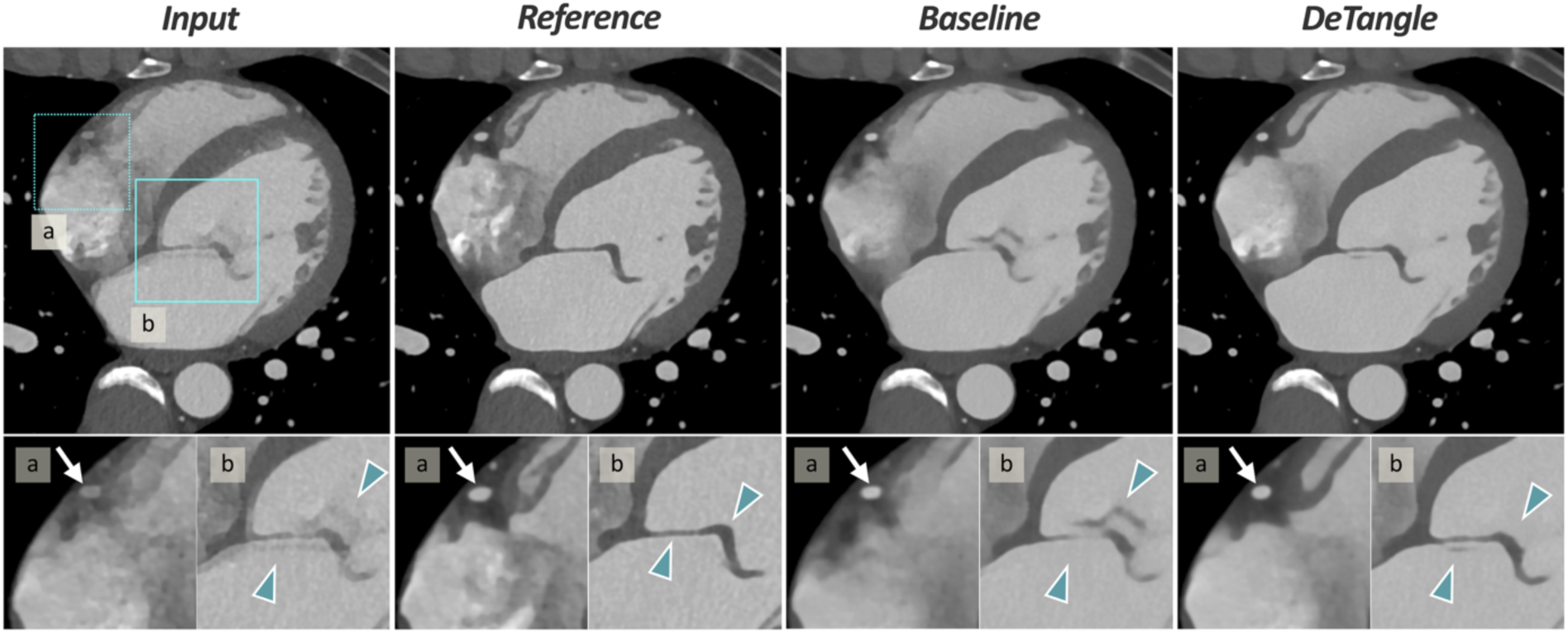
Improved cardiac computed tomography image quality with residual mitral valve blurring after DeTangle processing. Mid-diastolic cardiac computed tomography (CT) images with reconstruction starting 750 ms after the R wave of a man in his 40s show motion-blurred full reconstruction (MB-Full), motion-reduced half reconstruction (MR-Half), Baseline, and DeTangle (left to right), with magnified regions below. (a) Baseline improves delineation of the right coronary artery margins (white arrows). DeTangle better delineates the right ventricle, right atrium, and small vascular branches. (b) DeTangle improves mitral valve delineation (blue arrowheads) despite slight residual motion blur. Residual blur is also present in the MR-Half.

The DeTangle–Baseline SSIM difference increased with motion severity: +0.0016 (95% confidence interval [CI], 0.0012–0.0020), +0.0029 (95% CI, 0.0022–0.0035), and +0.0055 (95% CI, 0.0050–0.0060) for low-, intermediate-, and high-motion groups, respectively (P < .001).

### Visual image quality and local score reductions

The three-vessel mean coronary score was 3.3 with DeTangle versus 2.9 with Baseline (paired mean difference, +0.41 points; 95% CI, 0.35–0.46). Left ventricular myocardial scores were 4.2 versus 3.5 (common odds ratio [OR] for a higher score, 2.9; 95% CI, 2.3–3.7) (both Holm-adjusted P < .001). DeTangle also improved scores across all nine regions (all P < .001; Figure 6 and Supplemental Table S3).

**Figure 6.**
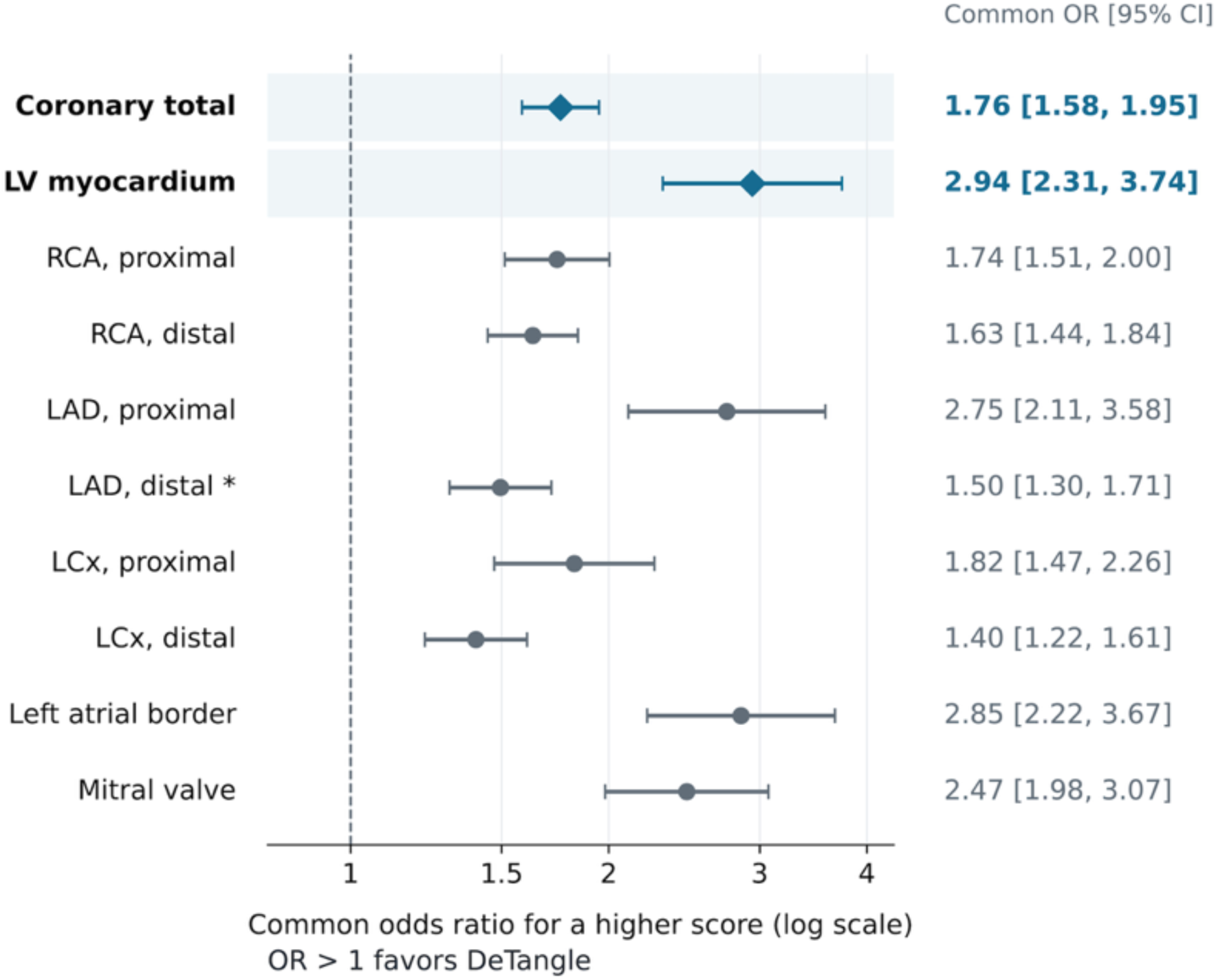
Visual image quality with DeTangle versus Baseline. Forest plot of common odds ratios (ORs) for higher visual scores with DeTangle versus Baseline. Diamonds indicate left ventricular (LV) myocardium and pooled coronary segments (“Coronary total”; segment-adjusted OR, 1.8; 95% confidence interval [CI], 1.6–1.9); circles indicate other regions. Bars show 95% CIs. Values right of OR = 1 on the logarithmic axis favor DeTangle. *The distal LAD was unevaluable because of occlusion in one patient, leaving 192 phases from 24 patients for that segment. RCA, right coronary artery; LAD, left anterior descending artery; LCx, left circumflex artery.

At least one primary score decreased with DeTangle in 9/200 phases (4.5%; Table 3). Across nine regions, at least one score decreased in 21/200 phases (10.5%). In one case, a distal RCA double contour appeared as a dissection-like double lumen after DeTangle processing (Supplemental Figure S3). Both readers favored DeTangle in the shared 100-phase subset (all Holm-adjusted P < .001), with interreader agreement detailed in Supplemental Table S4.

**Table 3.** Visual image quality scores for DeTangle compared with Baseline.

| **Outcome** | **Higher with DeTangle** | **Unchanged** | **Lower with DeTangle** |
|---|---|---|---|
| Three-vessel mean coronary score | 155 (77.5) | 39 (19.5) | 6 (3.0) |
| Left ventricular myocardial score | 103 (51.5) | 93 (46.5) | 4 (2.0) |

Values are numbers of cardiac phases out of 200 (percentages). Both primary outcome scores decreased in one phase; at least one decreased in 9/200 phases (4.5%).

### Quantitative image quality

DeTangle increased ERS and reduced ERD versus Baseline and MB-Full at all three sites (all Holm-adjusted P < .05, Table 4). Supplemental Figure S4 shows representative septal and coronary intensity profiles. Only LCA showed greater ERS gains and ERD reductions with increasing motion severity (Holm-adjusted P < .001 and P = .007, respectively; Supplemental Figure S5). DeTangle increased coronary attenuation, SNR, and CNR, whereas myocardial attenuation was similar (Supplemental Table S5).

**Table 4.** Edge rise slope and distance by image type.

| **Metric / Region** | **MB-Full** | **MR-Half** | **Baseline** | **DeTangle** | ***P* vs MB-Full** | ***P* vs MR-Half** | ***P* vs Baseline** |
|---|---|---|---|---|---|---|---|
| *ERS (HU/mm)* |  |  |  |  |  |  |  |
| RCA | 254.1 ± 123.9 | 440.0 ± 133.4 | 348.6 ± 164.2 | 401.8 ± 128.0 | <.001 | 0.004 | <.001 |
| LCA | 306.7 ± 120.6 | 432.6 ± 111.0 | 312.4 ± 152.4 | 411.5 ± 111.0 | <.001 | 0.053 | <.001 |
| IVS | 230.9 ± 134.0 | 374.6 ± 157.6 | 344.9 ± 149.1 | 383.2 ± 154.9 | <.001 | 0.32 | <.001 |
| *ERD (mm)* |  |  |  |  |  |  |  |
| RCA | 1.59 ± 0.44 | 1.24 ± 0.27 | 1.45 ± 0.39 | 1.28 ± 0.27 | <.001 | 0.25 | <.001 |
| LCA | 1.38 ± 0.30 | 1.16 ± 0.16 | 1.41 ± 0.33 | 1.18 ± 0.18 | <.001 | 0.78 | <.001 |
| IVS | 3.18 ± 2.16 | 1.39 ± 0.65 | 1.52 ± 0.86 | 1.28 ± 0.50 | <.001 | 0.04 | 0.01 |

Values are mean ± SD across 100 cardiac phases from 25 patients. Higher ERS and shorter ERD indicate sharper edges.
The displayed *P* values are for two-sided comparisons with DeTangle.
ERS, edge rise slope; ERD, edge rise distance; RCA, right coronary artery; LCA, left coronary artery, IVS, interventricular septum； MB-Full, motion-blurred full reconstruction; MR-Half, motion-reduced half reconstruction.

### Exploratory application to half reconstructions

DeTangle processing of MR-Half improved ERD and ERS at all three sites relative to both MR-Half and DeTangle-processed MB-Full (Supplemental Table S6).

### DM input perturbation and mixing experiments

Perturbing Teacher’s RealDM input reduced SSIM, with the greatest reduction occurring after map shuffling (mean difference, −0.14294; Supplemental Figure S6). Mixing 25%–50% RealDM with cDM slightly increased SSIM, whereas 75%–100% mixing reduced it (Supplemental Table S7).

### Inference time

DeTangle inference averaged 70 ms per slice.

## Discussion

This study evaluated whether DeTangle, which uses an internally estimated deformation map to guide image restoration, could reduce motion blur in coronary and noncoronary structures using only images from the target cardiac phase. Mean SSIM was higher with DeTangle (0.972) than with Baseline (0.968; P < .001). Compared with Baseline, DeTangle significantly improved coronary and left ventricular myocardial image-quality scores (both P < .001) and reduced the 10–90% edge rise distance in the interventricular septum and right and left coronary arteries (all P < .05). However, DeTangle did not reach the mean SSIM of Teacher, which directly used RealDM (0.976), and at least one primary outcome score was lower than with Baseline in 9/200 phases (4.5%).

Previous approaches used local coronary patches for direct image translation, multiphase processing, or motion estimation followed by projection-based compensation [15,16,23]. The latter estimated motion from 2.5D patches comprising three orthogonal planes [23], supporting the use of local through-plane information, which DeTangle instead obtains from three adjacent axial slices. Whole-heart approaches have also used multiple cardiac phases for image restoration or motion-compensated reconstruction [17,24,25]. Adapting motion-guided natural image deblurring [26,27], DeTangle jointly learns an internal scalar deformation-magnitude map and a restoration network conditioned on that map. Inference requires only three adjacent slices from the target phase, without additional cardiac phases, projection data, or externally prepared motion maps.

Consistent with previous applications of privileged information available only during training [28,29], DeTangle combined joint DMNet–Deblur optimization, motion-guided sampling, and Teacher distillation. Although DeTangle outperformed Baseline and LUPI-only, individual contributions remain unresolved. Teacher achieved the highest SSIM using RealDM, consistent with previous findings in super-resolution [28]. Shuffling RealDM markedly reduced SSIM, suggesting dependence on spatial organization. However, shuffling preserved the value distribution while disrupting both anatomical alignment and local smoothness. With Deblur weights fixed, complete replacement of cDM with RealDM reduced SSIM, whereas blending 25%–50% RealDM into cDM slightly improved it. Restoration may therefore depend on both the map representation and how the network has learned to use it [30]. Future studies should assess whether richer motion representations or uncertainty-aware maps improve restored image quality.

Motion-compensated reconstruction incorporates estimated motion fields and projection geometry into the reconstruction model [24,25,31]. DeTangle operates on reconstructed images without projection data or acquisition geometry at inference, offering a complementary image-based approach, although direct performance comparisons remain necessary. CT artifact patterns depend on projection angles, temporal sampling, and motion trajectories that may vary nonlinearly within the reconstruction window [32]. Differences between full and half reconstructions therefore motivated the exploratory half-input evaluation. Half-input processing further sharpened edges but did not establish an effective temporal resolution shorter than 66 ms. Full reconstructions may provide greater CT-number stability [33,34], which is relevant to quantitative applications such as radiomics. CT-number accuracy and radiomics feature reproducibility after deblurring require further validation.

DeTangle improved coronary and myocardial image quality, with additional benefits at the mitral valve and left atrial border. These findings suggest that postprocessing may aid visualization of myocardial and perivalvular structures in structural heart disease. Including image input and output, we estimate that offline processing of a 250-slice volume would take approximately 20 s. However, the observed local image-quality deterioration and potentially misleading artifacts (Supplemental Figure S3) highlight the need to improve restoration accuracy and suppress hallucinations before clinical use. Effects on lesion detection, diagnostic accuracy, and anatomical measurement accuracy remain unknown.

This study has limitations. First, the single-center cohort excluded arrhythmia and marked breath-hold failure. Training and the primary evaluation used 250-ms full reconstructions from a single energy-integrating detector CT scanner. Although half-input processing was explored, generalizability across institutions, scanners, and acquisition and reconstruction conditions remains unvalidated. Second, residual blur and differing temporal windows limit interpretation of reference agreement as motion-free recovery or 66-ms temporal resolution. Improvements in reference agreement may also reflect differences in noise and image texture. RealDM may be affected by registration errors, unrepresented motion variations, and reference-related information from MR-Half, limiting attribution of Teacher’s advantage solely to motion guidance. The physical accuracy of cDM remains unverified. Third, local 2.5D inputs and scalar maps may not fully capture spatial context or motion trajectories, warranting evaluation of 3D inputs and alternative slice counts. Finally, the visual and quantitative subsets emphasized systolic and mid-diastolic windows and contained proportionally fewer high-motion phases.

## Conclusion

DeTangle improved reference agreement and coronary and myocardial image quality relative to Baseline using only single-phase reconstructed CT images.

## Ethical Statement

This retrospective study was conducted in accordance with the Declaration of Helsinki guidelines, and the Research Ethics Committee of the National Cerebral and Cardiovascular Center approved this study (R20112-2), which waived the requirement for written informed consent.

## Data sharing statement

The data used in this study, including the training data, cannot be shared publicly to protect participant privacy. Non-image data will be shared upon reasonable request to the corresponding author, subject to ethics committee approval of the proposed research and the release of the requested materials. Training and evaluation code will be released on https://github.com/ttsynishii/DeTangle. Trained model weights will not be shared because they could enable reconstruction of training data.

## Declaration of generative AI and AI-assisted technologies in the manuscript preparation process

During manuscript preparation and analysis coding, the authors used ChatGPT (OpenAI, GPT-5.6, 6) for language editing, readability improvement, and assistance with code development and debugging. All AI-assisted outputs, including code suggestions, were reviewed, tested, revised, and edited by the corresponding author. The authors take full responsibility for the final manuscript, analyses, and conclusions. The ideas and interpretations presented in this work are solely those of the authors.

## Acknowledgments

The authors thank the CT technologists Yuji Matsuzaki, Shun Okuyama, and Masaki Sakurai for their support in image acquisition. The authors thank Editage (https://www.editage.jp/) for English-language editing. This work was supported by JSPS KAKENHI Grant Number 23K07149.

## List of abbreviations

cDM: network-predicted deformation-magnitude map
CT: computed tomography
DM: deformation-magnitude map
ERD: edge rise distance
ERS: edge rise slope
GEE: generalized estimating equation
MB-Full: motion-blurred full reconstruction
MR-Half: motion-reduced half reconstruction
RealDM: registration-derived deformation-magnitude map
SSIM: structural similarity index measure

