## Supplementary materials for "Deformation prior-guided deep learning for post hoc motion deblurring in single-phase cardiac computed tomography"

**Supplemental Materials**

### Supplemental Methods

#### *S1. Computed tomography (CT) acquisition and image preparation*

*S1.1. CT acquisition*

Cardiac CT was performed for indications including preoperative evaluation of valvular disease. Images were acquired using a 2 × 192 × 0.6-mm detector configuration, automatic tube potential selection (70–120 kV), and automatic exposure control (Supplemental Table S1). Contrast material containing 320 or 370 mg iodine/mL was injected over 10 s at an iodine delivery rate of 26.0 mg iodine/kg/s using a dual-head injector (Dual Shot GX 7; Nemoto Kyorindo). A second 10-s injection at half the initial concentration was followed by 30 mL of saline at the same flow rate. Scan timing was determined using a test injection and a region of interest (ROI) in the ascending aorta.

*S1.2. Reconstruction and reference image preparation*

Motion-blurred full reconstructions (MB-Full) and motion-reduced half reconstructions (MR-Half) were generated using ADMIRE strength 4, a Bv40 kernel, a 200-mm field of view, and 0.6-mm slice thickness and spacing. Post hoc denoising was applied only to MR-Half to reduce the noise difference associated with a shorter reconstruction window. The residual dense network algorithm described by Kobayashi et al. [S1] and used for coronary perivascular fat attenuation index assessment [S2] was applied using the same pretrained model as in previous mitral valve and quarter-dose coronary CT studies [S3, S4], which reported noise reductions of approximately 50–55%.

*S1.3. Deformation maps*

Within each 250-ms MB-Full window, the early restoration reference (MR-$Half_1$, 0–66 ms) was the fixed image, and the late reconstruction (MR-$Half_2$, 150–216 ms) was the moving image. RealDM was the voxel-wise magnitude of the 3D displacement vector estimated using SyN registration [S5] in ANTsPy [S6]. Displacement values were stored in 0.1-mm units. The phase-level motion score was the 95th percentile of RealDM values exceeding two stored units (0.2 mm) across slices. Tertiles of phase-level motion scores across the 424 internal test phases defined low (<2.6 mm), intermediate (2.6 to <4.2 mm), and high (≥4.2 mm) motion.

The first registration stage used SyN with 50, 30, 20, 10, and 5 iterations across the five resolution levels. The second stage used SyN with a cross-correlation (CC) similarity metric and a neighborhood radius of two voxels, with 40, 20, and five iterations across three resolution levels, and was initialized with the deformation transform from the first stage.

*S1.4. Reference-agreement metrics*

SSIM, PSNR, and LPIPS were calculated over the entire image for each slice and averaged across slices within each cardiac phase. For all three metrics, predicted and reference images were clipped to −1024 to 3071 HU and linearly scaled to [0, 1]. The SSIM and PSNR used a data range of 1. For AlexNet-based LPIPS, each normalized grayscale slice was replicated across three channels.

### *S2. Network architecture and training*

*S2.1. Architecture and spatial correspondence*

All four configurations used the same Swin-Conv-UNet (SCUNet)-based Deblur architecture, except for the number of input channels. Baseline and learning using privileged information (LUPI)-only received only three adjacent MB-Full slices. Teacher and DeTangle received the same three image channels and a fourth channel containing RealDM and cDM, respectively. Deblur used 96 × 96-pixel training patches. DMNet received a 128 × 128-pixel patch enclosing the Deblur patch and generated a 64 × 64-pixel cDM. The corresponding 48 × 48-pixel region was cropped and bilinearly upsampled to 96 × 96 pixels before concatenation with the image channels. The recorded SCUNet block configurations were 2, 2, 2, 3, 2, 2, and 2 for Deblur and 1, 2, 2, 3, 2, 2, and 1 for DMNet, respectively.

*S2.2. Training stages and objective*

DMNet pretraining was followed by adaptation with fixed Deblur and joint optimization. DeTangle training was configured for 210 epochs: an initial 10-epoch period followed by a 200-epoch DM-based patch-sampling curriculum. For all four configurations, each training epoch sampled one patch from every training image. Baseline randomly selected patch locations within each image. LUPI-only, Teacher, and DeTangle used the same RealDM-based curriculum for selecting patch locations. DeTangle traversed the training images in a shuffled order. During the initial period, the low, high, and uniformly random sampling probabilities were 0.25, 0.75, and 0, respectively. Over the first half of the subsequent curriculum, these probabilities changed linearly to 0, 0.50, and 0.50, remained at these values until 80% progress, and then changed to 0, 0.25, and 0.75. Rotation or flip augmentation was not applied during training. Both the Deblur and DMNet networks used separate Adam optimizers. Learning rates remained constant at $5 \times 10^{-5}$ for Deblur and $5 \times 10^{-6}$ for DMNet. DMNet was frozen for the first five epochs of joint training and was subsequently updated every eighth mini-batch; gradient clipping was disabled.

Each validation pass evaluated one 96 × 96-pixel patch per slice across the validation set. Patch locations were resampled at each validation pass using fixed sampling probabilities of 25% high motion and 75% uniform random. The checkpoint with the highest mean validation SSIM was selected.

The joint objective combined a Charbonnier reconstruction loss, a z-direction residual-consistency term, an output-distillation term, and cDM anchor and range terms with weights of 1, 0.1, 0.5, 0.03, and 0.05, respectively. The consistency term compared the prediction-minus-reference residuals in adjacent slices after 2 × 2 average pooling, with gradients stopped through the neighboring prediction. The anchor term combined absolute cDM–RealDM differences with a relative term, $0.3 \times \log[1 + |\text{cDM} - \text{RealDM}|/(|\text{RealDM}| + 0.001)]$, in normalized DM units. The range term penalized values that fell below 0 or above 1, and both regularizers used HU window-based spatial weights.

The joint-training configuration applied final-resolution output distillation during epochs 1–200, and disabled it during epochs 201–210. The output-distillation loss (OutputKD) used a Charbonnier penalty averaged over pixels for which Teacher's absolute error was at least 2 HU lower than the student's; if no pixels qualified, the unmasked mean was used. LUPI-only used output, feature, and attention distillation from Teacher. In contrast, DeTangle used only final-output distillation, allowing Deblur to learn intermediate features and attention patterns suited to cDM-guided restoration.

| **Setting** | **Recorded DeTangle value** |
|---|---|
| Deblur learning rate | $5 \times 10^{-5}$ |
| DMNet learning rate | 0.1 × Deblur learning rate |
| Batch size / gradient accumulation | 8 / 4 |
| DMNet updates | Every eighth mini-batch after the first five epochs of joint training (frozen) |
| z-residual downsampling factor | 2 |
| Feature / attention distillation weights | 0 / 0 |
| DM floor / maximum clipping value | 2 / 130 stored units |
| DMNet HU range | −900 to 3000 HU, with window mask |
| Drop-path rate | 0.01 |
| Numerical precision | Automatic mixed precision |

DM, deformation-magnitude map; DMNet, deformation-magnitude map prediction network; HU, Hounsfield units

#### *S3. Selection of visual and quantitative evaluation phases*

*S3.1. Phase selection for Reader 1 (200 phases)*

The systolic reconstruction start times were 300, 350, and 400 ms after the R wave. For three patients with shorter RR intervals, these were adjusted to 250, 300, and 350 ms because the reconstruction window starting at 400 ms overlapped with the subsequent diastolic period. Mid-diastolic candidates were selected from saved phases within an RR-relative window, typically RR−350 to RR−200 ms, or RR−450 to RR−300 ms when the RR-equivalent value exceeded 1100 ms. For each patient, the two out-of-window phases were drawn uniformly from eligible pairs separated by at least 250 ms.

*S3.2. Phase selection for Reader 2 and quantitative evaluation (100 phases)*

The fixed phases were the 300-ms phase and the middle of the three mid-diastolic candidates in the ordered reconstruction time. One additional phase was drawn uniformly from the remaining systolic candidates for 12 randomly assigned patients and from the remaining mid-diastolic candidates for the other 13 patients. The fourth phase was drawn uniformly from the two previously selected out-of-window phases.

#### *S4. Visual scoring and missing observations*

The first reader was a board-certified radiologist with 19 years' experience (T.N.). The second was a board-certified radiological technologist with six years of cardiac CT experience (T.S.). Reader 1 scored all 200 phases and positioned regions of interest (ROIs) for the 100-phase subset used in quantitative evaluation. Reader 2 scored the same 100-phase subset. The nine scored regions were the proximal and distal segments of the RCA, LAD, and left circumflex artery, together with the left ventricular myocardium, left atrial border, and mitral valve. Higher scores indicated better image quality.

| Score | Image quality criterion |
|---|---|
| 1 | Severe disruption of the anatomical structure. |
| 2 | The structure can be traced, but its boundaries are indistinct with partial disruption. |
| 3 | The structure can be traced, but its edges show blurring or double contours. |
| 4 | Slight residual edge blurring. |
| 5 | Clear, well-defined edges. |

The score for each coronary artery was the mean of the available segment scores. The three-vessel mean coronary score was calculated as the average of the RCA, LAD, and left circumflex artery scores. Distal LAD occlusion rendered the segment unevaluable for one patient across all phases in both models. The proximal LAD score was used to represent the LAD in the three-vessel mean coronary score for this patient. Improvement, no change, and deterioration were defined according to the sign of the paired DeTangle–Baseline score difference. A phase was considered to have local deterioration if any evaluable region had a lower DeTangle score. For inter-reader agreement, paired scores were pooled across Baseline and DeTangle within each region. Pairs were excluded if either reader marked the region as unevaluable.

### *S5. Quantitative measurements and aggregation*

ROI placement was primarily based on the MR-Half reference images, with all corresponding image types reviewed. Circular ROIs were repositioned when necessary to avoid artifacts, and line ROI positions were corrected when displaced from the intended anatomical target. The resulting ROIs were therefore image-specific, with coordinates adjusted as needed for each image type.

#### *S5.1. Edge metrics*

Reader 1 placed image-specific line ROIs. On axial images, coronary line ROIs were oriented perpendicularly to the local vessel course at the proximal and mid RCA, proximal LAD, and proximal left circumflex artery. For each phase and image type, the septal summary averaged measurements from the three lines. The RCA summary combined the proximal and mid-RCA lines, and the LCA summary combined the proximal LAD and left circumflex artery lines. Each coronary summary averaged four measurements from both edges of the two lines. ERS and ERD were averaged separately. The 10% and 90% crossings of the local intensity increase were determined using linear interpolation. ERD was defined as the distance between these crossings, and ERS as the intensity increase divided by ERD.

#### *S5.2. Attenuation, noise, SNR, and CNR*

Three circular ROIs were placed in each of the ascending aorta, left ventricular blood pool, and left ventricular myocardium, with diameters of 20, 15, and 8 mm, respectively, while avoiding severe artifacts. For each anatomical region, attenuation was calculated as the mean of the three ROI mean CT values, and noise as the mean of their within-ROI standard deviations (SDs). Coronary attenuation was calculated as the average of the mean CT values in three circular ROIs (1.5–2.0 mm in diameter) placed in the proximal LAD, proximal left circumflex artery, and proximal RCA.

SNR was defined as the regional mean attenuation divided by regional noise. CNR was defined as the signed difference between target and myocardial attenuation divided by the mean of their noise values. Coronary SNR used ascending aortic noise from the same phase and image type. Coronary CNR was defined as the difference between coronary and myocardial attenuation divided by the mean of the ascending aortic and myocardial noise values. Ratios were calculated for each phase before averaging. CT attenuation, noise, SNR, and CNR measurements are presented in Supplemental Table S5.

***S6. Exploratory application to half-reconstructions***

DeTangle was applied to denoised 66-ms MR-Half images from the same 100 phases used for quantitative evaluation (Supplemental Methods S1.2 and S3.2). ROI placement, edge measurements, and phase-level aggregation followed the procedures described in S5, using image-specific ROIs that were reviewed and adjusted for half-input outputs. These outputs were compared with MR-Half and the corresponding DeTangle outputs generated from full-reconstruction inputs (Supplemental Table S6).

***S7. DM input perturbation and mixing experiments***

The Teacher model's RealDM input was downsampled to half resolution, scaled by 0.25 or 0.5, set to zero, or shuffled in separate experiments (Supplemental Figure S6). For shuffling, all pixel values within each 2D RealDM slice were randomly permuted, preserving the value distribution while disrupting both the spatial correspondence with the CT image and the local smoothness. Shuffling was applied at the original resolution and amplitude ($r = 1$, $\gamma = 1$), with one permutation per slice.

For mixing, RealDM was converted into the representation used by DeTangle's Deblur network. The mixed map was defined as $(1 - \alpha)$cDM + $\alpha$RealDM, where $\alpha$ represented the RealDM fraction. The unmixed cDM comparator ($\alpha = 0$) came from the same inference run. The RealDM fractions ($\alpha$) were 0.25, 0.50, 0.75, and 1.00, corresponding to 25%, 50%, 75%, and 100% RealDM, respectively.

***S8. Additional statistical analyses***

*S8.1. Reference agreement and quantitative outcomes*

For the main four-image comparisons, reference agreement and quantitative metrics were analyzed using Gaussian GEEs, with Holm adjustment across six pairwise comparisons per metric. The DeTangle–Baseline SSIM difference was tested for an unadjusted linear trend across ordered motion groups. Trends in DeTangle–Baseline differences in 10%–90% edge rise slope and edge rise distance were assessed using image-type × ordered-motion-group interactions, with Holm adjustment across the six outcomes.

*S8.2. Visual outcomes and interreader agreement*

Visual comparisons used Gaussian GEEs for paired coronary mean score differences and cumulative logit ordinal GEEs for myocardial and regional scores. Holm adjustment was applied separately to the two principal outcomes and the nine regional outcomes within each reader and evaluation set. A supportive analysis pooled the six coronary segments using a cumulative logit ordinal GEE with adjustment for segment to estimate the common odds ratio for a higher score with DeTangle. Interreader agreement was assessed separately for each region using linearly weighted Cohen $\kappa$ in the shared 100-phase subset.

*S8.3. Exploratory perturbation and mixing analyses*

Teacher perturbation effects were analyzed as phase-level paired SSIM differences relative to Teacher with its original RealDM input, using separate intercept-only Gaussian GEEs for each perturbation. Two-sided tests used Holm adjustment across the five perturbation comparisons. Mixing experiments used a Gaussian GEE with categorical RealDM fraction, with Holm adjustment across four comparisons against unmixed cDM.

*S8.4. Confidence intervals*

GEE-based 95% confidence intervals (CIs) and two-sided hypothesis tests used a t-approximation with degrees of freedom equal to the number of patient clusters contributing to the analysis minus one. All GEE analyses used the variance estimator specified in the main text. Odds ratio CIs were calculated on the log-odds scale and exponentiated.

Motion-stratified mean paired differences were estimated using separate intercept-only Gaussian GEEs for each motion stratum. Means were phase-weighted. For the low-, intermediate-, and high-motion strata, the contributing patient counts were 24/25/24 for SSIM and 22/21/15 for edge metrics, respectively; Teacher perturbation analyses included 25 patients. Motion-stratified CIs describe mean gains within each stratum, rather than the coefficients used to test trends across strata. CIs for weighted κ were estimated using patient-cluster percentile bootstrap resampling with 20,000 replicates, retaining all paired phases and both image types within each sampled patient.

### Supplementary Tables

#### Table S1. Cardiac CT acquisition and reconstruction settings for MB-Full and MR-Half

| CT Parameters | MB-Full image | MR-Half image |
|---|---|---|
| *Scan parameters* | | |
| Scan mode | Retrospective ECG-gated helical scan | |
| Collimation (mm) | 2×192×0.6 | |
| Rotation speed (ms) | 250 | |
| Tube potential (kV) | 70, 80, 90, 100, 120 | |
| AEC | CARE Dose 4D, CARE kV | |
| AEC setting | Quality ref. mAs/rot. 270 with Ref. kV 120 | |
| Pitch | 0.15-0.34 | |
| Contrast injection rate (mgI/kg/sec) | 26 | |
| *Reconstruction parameters* | | |
| Reconstruction mode | Full (250 ms) | Half (66 ms) |
| Slice thickness/interval (mm) | 0.6/0.6 | |
| Reconstruction phases | 4D, 50-ms steps across cardiac phase | |
| Kernels | Bv40 / ADMIRE strength 4 | |
| Field of view (mm) | 200 | |

Note: MB-Full (motion-blurred full reconstruction, 250-ms window) and MR-Half (motion-reduced half reconstruction, 66-ms window) images were generated from the same retrospective ECG-gated helical acquisition. ECG, electrocardiogram; AEC, automatic exposure control.

**Table S2. Model configuration, training, and inference inputs**

| Model | Deblur input | Training patch-location sampling | Additional use of RealDM during training | Distillation from Teacher | External inputs at inference |
|---|---|---|---|---|---|
| Baseline | Three adjacent MB-Full slices | Random location within each image | — | None | MB-Full |
| LUPI-only | Three adjacent MB-Full slices | Shared RealDM-based curriculum | — | Output, feature, and attention | MB-Full |
| DeTangle | Three adjacent MB-Full slices and internally predicted cDM | Shared RealDM-based curriculum | Supervises cDM during joint training | Final output only | MB-Full |
| Teacher | Three adjacent MB-Full slices and RealDM | Shared RealDM-based curriculum | Direct input to Deblur | None (distillation source) | MB-Full and RealDM |

Note: Each training epoch used one patch from each training image for all four configurations. The Deblur architecture is shared apart from the input channels. MB-Full, motion-blurred full reconstruction; RealDM, registration-derived deformation-magnitude map; cDM, deformation-magnitude map predicted using DMNet.

**Table S3. Visual image quality scores for DeTangle compared with Baseline**

| Outcome | Paired phases | Baseline | DeTangle | OR (95%CI) | Holm-adjusted *P* |
|---|---|---|---|---|---|
| *Primary outcomes* | | | | | |
| Coronary total* | 200 | 2.9 ± 1.1 | 3.3 ± 1.0 | 1.8 (1.6–1.9) | – |
| Left ventricular myocardium | 200 | 3.5 ± 1.3 | 4.2 ± 1.0 | 2.9 (2.3–3.7) | <.001 |
| *Exploratory regional outcomes* | | | | | |
| Proximal RCA | 200 | 2.8 ± 1.3 | 3.2 ± 1.3 | 1.7 (1.5–2.0) | <.001 |
| Distal RCA | 200 | 2.7 ± 1.3 | 3.0 ± 1.3 | 1.6 (1.4–1.8) | <.001 |
| Proximal LAD | 200 | 3.3 ± 1.5 | 4.0 ± 1.1 | 2.7 (2.1–3.6) | <.001 |
| Distal LAD | 192 | 3.1 ± 1.3 | 3.4 ± 1.2 | 1.5 (1.3–1.7) | <.001 |
| Proximal LCx | 200 | 3.0 ± 1.4 | 3.4 ± 1.2 | 1.8 (1.5–2.3) | <.001 |
| Distal LCx | 200 | 2.7 ± 1.3 | 2.9 ± 1.3 | 1.4 (1.2–1.6) | <.001 |
| Left atrial border | 200 | 3.6 ± 1.1 | 4.1 ± 0.9 | 2.9 (2.2–3.7) | <.001 |
| Mitral valve | 200 | 3.0 ± 1.2 | 3.6 ± 1.2 | 2.5 (2.0–3.1) | <.001 |

Note: Scores are mean ± SD. ORs indicate higher DeTangle scores.
* In the Coronary total row, the descriptive scores summarize the primary three-vessel mean score (paired mean difference +0.41 points, 95% CI 0.35–0.46, Holm-adjusted P < .001). The OR of 1.8 (95% CI 1.6–1.9) comes from a separate supportive analysis pooling six segments with adjustment for segment.
CI, confidence interval; OR, odds ratio; RCA, right coronary artery; LAD, left anterior descending artery; LCx, left circumflex artery.

**Table S4. Visual image quality and inter-reader agreement in the shared 100 Phases**

**S4A. Reader-specific effects**

| Outcome | Reader 1 (95% CI) | Reader 2 (95% CI) |
|---|---|---|
| Coronary total, OR | 1.7 (1.5–1.9) | 2.0 (1.8–2.4) |
| Three-vessel mean coronary score, mean difference (points) | 0.37 (0.29–0.44) | 0.50 (0.41–0.59) |
| Left ventricular myocardium, OR | 3.0 (2.2–4.2) | 3.0 (2.2–4.1) |

Note: Both readers assessed the same 100 phases in the 25 patients. The mean differences were defined as DeTangle minus baseline. ORs indicate higher DeTangle scores. Both primary outcomes were Holm-adjusted. *$P$* <.001 for each reader. The coronary total was the segment-adjusted OR pooled across the six coronary segments.
OR, odds ratio; CI, confidence interval

**S4B. Inter-reader agreement**

| Region | Paired scores | Weighted κ (95% CI) |
|---|---|---|
| Proximal RCA | 200 | 0.71 (0.63–0.77) |
| Distal RCA | 200 | 0.76 (0.68–0.83) |
| Proximal LAD | 200 | 0.74 (0.66–0.79) |
| Distal LAD | 192 | 0.80 (0.72–0.87) |
| Proximal LCx | 200 | 0.75 (0.69–0.81) |
| Distal LCx | 200 | 0.71 (0.59–0.80) |
| Left ventricular myocardium | 200 | 0.69 (0.60–0.76) |
| Left atrial border | 200 | 0.62 (0.55–0.68) |
| Mitral valve | 200 | 0.67 (0.57–0.74) |

Note: Linearly weighted Cohen κ pools Baseline and DeTangle score pairs. Four phases from one patient were excluded from the distal LAD, yielding 192 paired scores.
CI, confidence interval; OR, odds ratio; RCA, right coronary artery; LAD, left anterior descending artery; LCx, left circumflex artery.

**Table S5. CT attenuation, noise, SNR, and CNR**

| Metric / Region | MB-Full (input) | MR-Half (reference) | Baseline | DeTangle | *P* vs MB-Full | *P* vs MR-Half | *P* vs Baseline |
|---|---|---|---|---|---|---|---|
| *CT attenuation (HU)* | | | | | | | |
| Ascending aorta | 611 ± 132 | 613 ± 131 | 610 ± 131 | 611 ± 131 | 0.35 | 0.35 | <.001 |
| Myocardium | 121 ± 33 | 117 ± 22 | 118 ± 22 | 117 ± 22 | 0.36 | 0.68 | 0.65 |
| LV blood pool | 630 ± 146 | 631 ± 145 | 629 ± 147 | 632 ± 145 | 0.043 | 0.63 | 0.01 |
| Coronary arteries | 434 ± 147 | 564 ± 134 | 463 ± 174 | 551 ± 118 | <.001 | 0.08 | <.001 |
| *Image noise (HU)* | | | | | | | |
| Ascending aorta | 20 ± 3 | 20 ± 4 | 11 ± 2 | 10 ± 2 | <.001 | <.001 | <.001 |
| Myocardium | 21 ± 5 | 19 ± 5 | 12 ± 4 | 10 ± 3 | <.001 | <.001 | <.001 |
| LV blood pool | 23 ± 4 | 22 ± 4 | 14 ± 8 | 12 ± 2 | <.001 | <.001 | 0.02 |
| *SNR* | | | | | | | |
| Ascending aorta | 31.5 ± 6.4 | 31.2 ± 7.3 | 58.2 ± 11.2 | 59.7 ± 11.4 | <.001 | <.001 | <.001 |
| Myocardium | 5.8 ± 1.2 | 6.2 ± 1.2 | 10.3 ± 2.3 | 11.8 ± 2.3 | <.001 | <.001 | <.001 |
| LV blood pool | 27.6 ± 6.6 | 28.6 ± 6.9 | 49.7 ± 12.8 | 54.3 ± 11.1 | <.001 | <.001 | <.001 |
| Coronary arteries | 22.5 ± 7.7 | 28.7 ± 7.4 | 44.3 ± 16.5 | 53.9 ± 11.3 | <.001 | <.001 | <.001 |
| *CNR* | | | | | | | |
| Ascending aorta | 24.3 ± 5.8 | 25.6 ± 6.5 | 44.3 ± 10.4 | 48.5 ± 10.7 | <.001 | <.001 | <.001 |
| LV blood pool | 23.1 ± 6.0 | 25.0 ± 6.7 | 41.5 ± 11.7 | 47.2 ± 11.1 | <.001 | <.001 | <.001 |
| Coronary arteries | 15.7 ± 7.2 | 23.1 ± 6.6 | 31.3 ± 15.3 | 42.8 ± 10.6 | <.001 | <.001 | <.001 |

Note: MR-Half images underwent deep learning denoising. The ascending aortic noise was lower in both restoration outputs (baseline, 11 HU; DeTangle, 10 HU) than in MB-Full or denoised MR-Half (both 20 HU), contributing to a higher SNR and CNR. P-values compare DeTangle with each indicated image type and were Holm-adjusted across all six image pairs for each regional metric. Coronary SNR uses the ascending aortic noise, and coronary CNR uses the mean of the ascending aortic and myocardial noise.

HU, Hounsfield unit; MB-Full, motion-blurred full reconstruction; MR-Half, motion-reduced half reconstruction; LV, left ventricle; SNR, signal-to-noise ratio; CNR, contrast-to-noise ratio.

**Table S6. Edge rise slope and distance after DeTangle processing of half-reconstructions**

| Metric / Region | MR-Half | DeTangle | DeTangle (MR-Half input) | *P* vs MR-Half | *P* vs DeTangle |
|---|---|---|---|---|---|
| *ERS (HU/mm)* | | | | | |
| RCA | 440.0 ± 133.4 | 401.8 ± 128.0 | 493.6 ± 139.7 | <.001 | <.001 |
| LCA | 432.6 ± 111.0 | 411.5 ± 111.0 | 466.3 ± 127.3 | <.001 | <.001 |
| IVS | 374.6 ± 157.6 | 383.2 ± 154.9 | 431.5 ± 170.4 | <.001 | <.001 |
| *ERD (mm)* | | | | | |
| RCA | 1.24 ± 0.27 | 1.28 ± 0.27 | 1.16 ± 0.21 | 0.005 | <.001 |
| LCA | 1.16 ± 0.16 | 1.18 ± 0.18 | 1.08 ± 0.14 | <.001 | <.001 |
| IVS | 1.39 ± 0.65 | 1.28 ± 0.50 | 1.09 ± 0.30 | <.001 | <.001 |

Note: Values are means ± SD across the same 100 phases from 25 patients. MR-Half denotes denoised 66-ms half reconstructions. DeTangle and DeTangle (MR-Half input) denote the outputs of the full and half reconstruction inputs, respectively. Displayed P-values compare DeTangle (MR-Half input) with the indicated image types. Paired differences were analyzed using patient-clustered Gaussian GEE with independent working correlation and Mancl–DeRouen variance correction. Holm adjustment was applied across six outcomes (ERS and ERD in the RCA, LCA, and IVS) separately for each comparator (MR-Half and full-input DeTangle).

MR-Half, motion-reduced half reconstruction; SD, standard deviation; GEE, generalized estimating equation; ERS, edge rise slope; ERD, edge rise distance; RCA, right coronary artery; LCA, left coronary artery; IVS, interventricular septum.

**Table S7. Effect of mixing cDM and RealDM on SSIM**

| RealDM fraction | Mean ΔSSIM | 95% CI | Holm-adjusted *P* |
|---|---|---|---|
| 25% | 0.00023 | +0.00018 to +0.00028 | <.001 |
| 50% | 0.00025 | +0.00013 to +0.00038 | <.001 |
| 75% | −0.00036 | −0.00063 to −0.00009 | 0.01 |
| 100% | −0.00227 | −0.00280 to −0.00173 | <.001 |

Note: ΔSSIM is the mean paired phase-level difference relative to unmixed cDM (0% RealDM) from the same inference run, across 424 phases. DeTangle's restoration network was fixed.
SSIM, structural similarity index; cDM, predicted deformation-magnitude map; CI, confidence interval

### Supplementary Figures

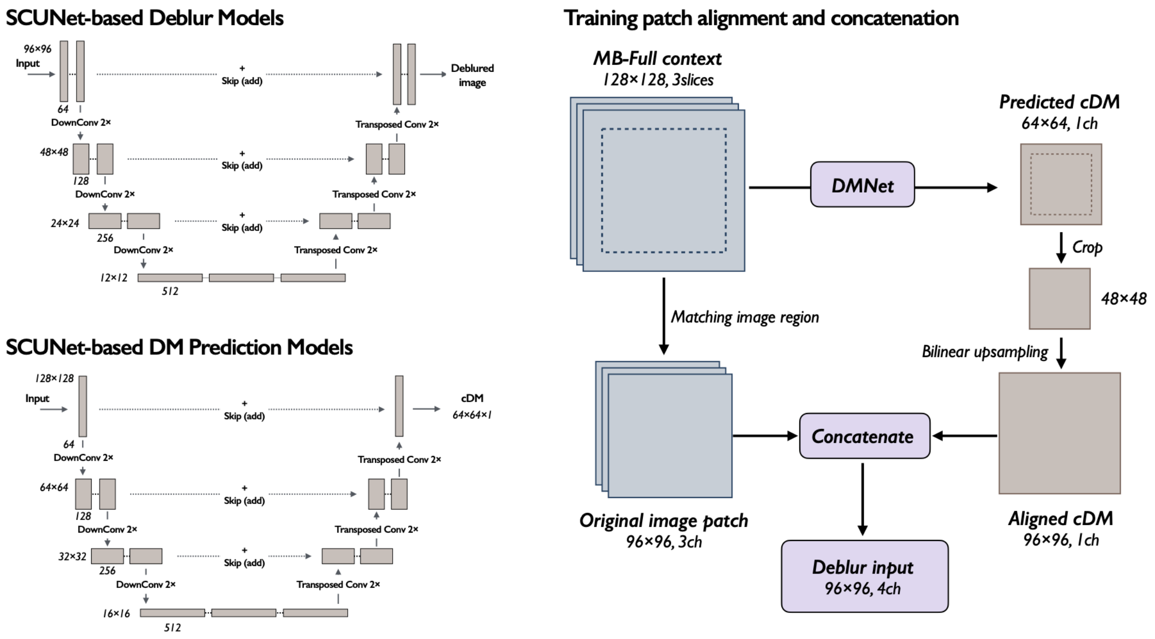


**Figure S1. DeTangle network architecture and training patch alignment**
Left, SCUNet-based architectures of the deblurring network (Deblur, top) and deformation-magnitude map prediction network (DMNet, bottom). Right: Dashed boxes indicate spatially corresponding regions in the motion-blurred full-reconstruction CT input (MB-Full) and predicted deformation-magnitude map (cDM). The aligned cDM was concatenated with three image channels to form the Deblur training input. Implementation details are provided in Supplemental Methods S2.1. ch, channels; cDM, predicted deformation-magnitude map; CT, computed tomography; DMNet, deformation-magnitude map prediction network; MB-Full, motion-blurred full reconstruction; SCUNet, Swin-Conv-UNet.

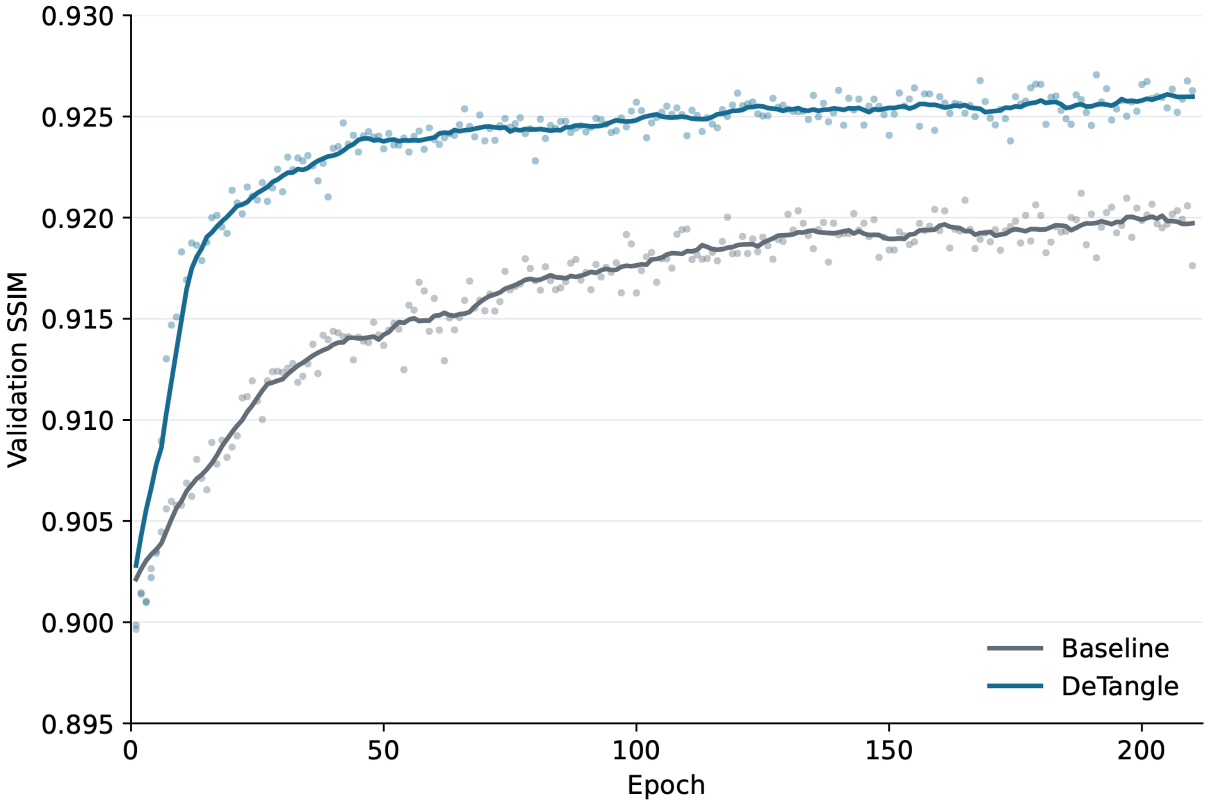


**Figure S2. Learning curves for Baseline and DeTangle.**

Points represent the mean SSIM of validation patches at each training epoch for Baseline (gray) and DeTangle (blue). Solid lines: 11-epoch moving averages, calculated using available epochs at the boundaries. All 210 training epochs are shown.

SSIM, structural similarity index

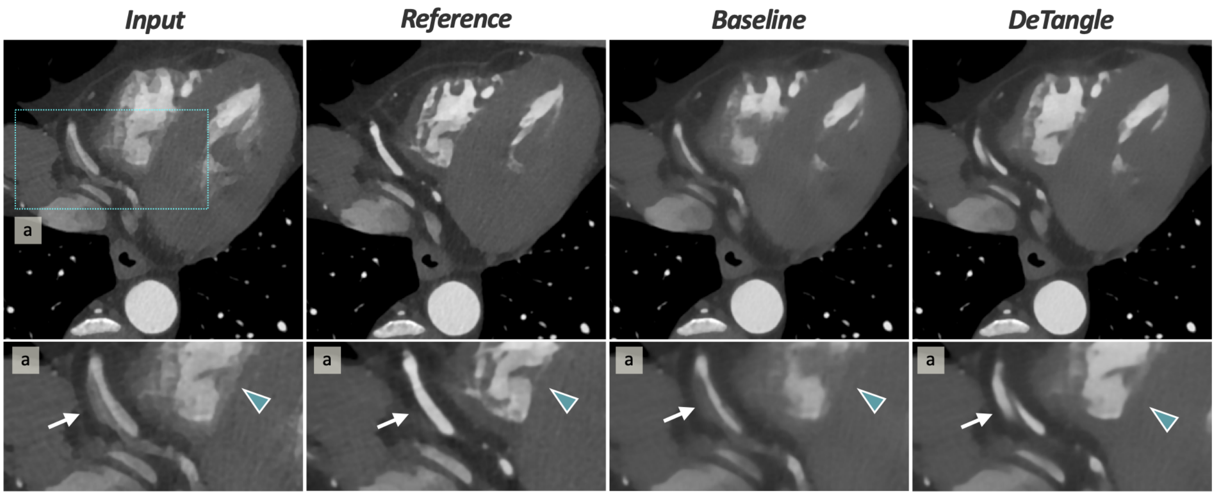


**Figure S3. Local image degradation after DeTangle processing.**
Axial cardiac CT images of a male in their 40s at end-systole (300 ms after the R wave) show MB-Full, MR-Half, Baseline output, and DeTangle output from left to right. The bottom row shows magnified views of region a. The distal right coronary artery (white arrows) exhibits a double contour in the input image. The Baseline output shows a continuous lumen with irregular margins, whereas the DeTangle output exhibits an apparent double lumen mimicking coronary dissection, an artifact absent from the reference image. Right ventricular trabeculae (blue arrowheads), obscured by artifacts in the Baseline output, are better delineated in the DeTangle output.

CT, computed tomography; MB-Full, motion-blurred full reconstruction; MR-Half, motion-reduced half reconstruction.

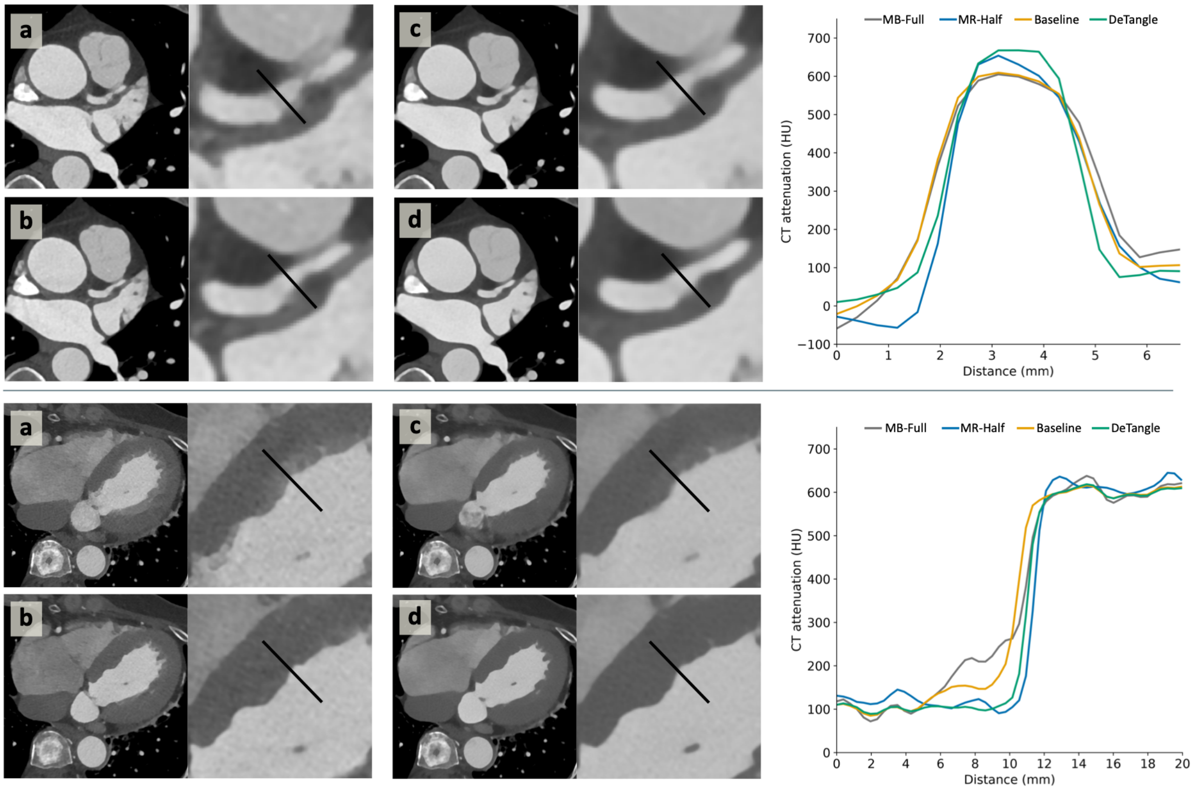


**Figure S4. LAD and interventricular septal images with corresponding CT attenuation profiles.** The upper and lower blocks show the LAD and interventricular septum, respectively. In each block, panels (a)–(d) show MB-Full, MR-Half, Baseline, and DeTangle output, respectively. Each panel contains an overview CT image and a corresponding magnified view. The plots to the right show overlaid CT attenuation profiles, with black lines in the magnified images indicating sampling locations. The horizontal and vertical axes represent distance (mm) and CT attenuation (HU), respectively.

For the LAD, the mean left- and right-edge 10%–90% ERS and ERD, respectively, were 323.5 HU/mm and 1.34 mm for MB-Full, 436.4 HU/mm and 1.25 mm for MR-Half, 371.8 HU/mm and 1.18 mm for Baseline, and 494.4 HU/mm and 1.00 mm for DeTangle. Corresponding values for the interventricular septum were 73.8 HU/mm and 5.43 mm, 370.1 HU/mm and 1.11 mm, 181.9 HU/mm and 2.20 mm, and 287.8 HU/mm and 1.39 mm, respectively.

CT, computed tomography; ERD, edge rise distance; ERS, edge rise slope; HU, Hounsfield units; LAD, left anterior descending coronary artery; MB-Full, motion-blurred full reconstruction; MR-Half, motion-reduced half reconstruction.

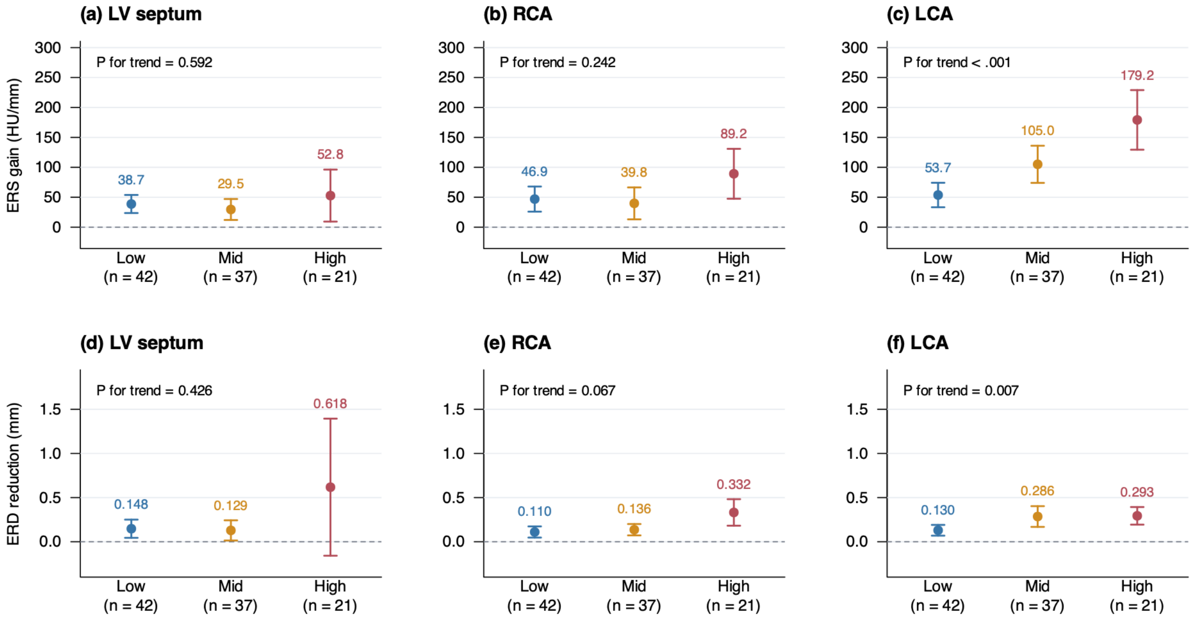


**Figure S5. Motion-stratified improvements in edge sharpness with DeTangle.**
Panels (a)–(c) show ERS gains (DeTangle minus Baseline), and panels (d)–(f) show ERD reductions (Baseline minus DeTangle) for the interventricular septum, RCA, and LCA, respectively. Both metrics were measured over the 10%–90% intensity transition interval, with positive values favoring DeTangle. Points represent mean paired differences in the low- (42 phases), intermediate- (37 phases), and high-motion (21 phases) groups. Error bars indicate unadjusted 95% CIs. Linear trends across ordered motion groups were tested with Holm's adjustment across six outcomes. Improvements increased with motion severity for LCA ERS (adjusted $P < .001$) and ERD (adjusted $P = .007$).

CI, confidence interval; ERD, edge rise distance; ERS, edge rise slope; LCA, left coronary artery; RCA, right coronary artery

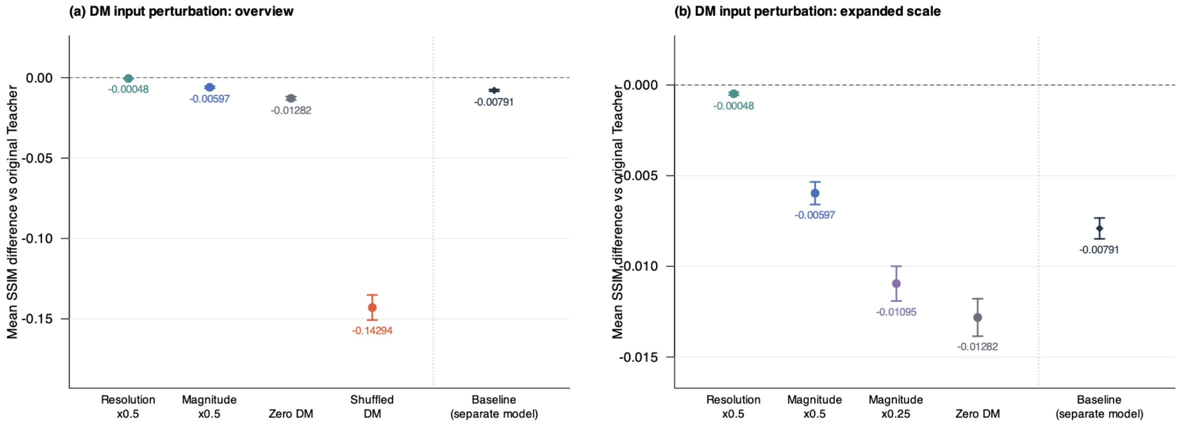


**Figure S6. Effect of perturbing the Teacher model's deformation-magnitude map input.**
Points represent the mean paired ΔSSIM across 424 cardiac phases from 25 patients, calculated as the SSIM for each condition or model minus that of the Teacher model using its original registration-derived RealDM. The horizontal dashed line indicates ΔSSIM = 0; more negative values indicate greater loss of agreement with the reference images. Panel (a) shows spatial shuffling of RealDM, which produces the largest decrease. Panel (b) uses an expanded vertical scale, excludes the shuffling condition, and includes RealDM scaled to 25% of its original magnitude. Diamonds represent Baseline, a separately trained image-only model, in both panels. All five RealDM perturbations reduced SSIM relative to Teacher with its original RealDM input (all two-sided Holm-adjusted $P < .001$; patient-clustered Gaussian GEEs). Baseline was shown for reference and was not included in these five comparisons.

ΔSSIM, difference in structural similarity index; RealDM, registration-derived deformation-magnitude map; SSIM, structural similarity index
